# The value of freedom: the introduction of option freedom in health-related capability wellbeing measurement

**DOI:** 10.1101/2022.10.05.22280720

**Authors:** Jasper Ubels, Karla Hernandez-Villafuerte, Michael Schlander

**Author notes:** **Corresponding Author:** Jasper Ubels. E-Mail other authors: Michael Schlander, Karla Hernández-Villafuerte.

## Abstract

The capability approach has been used to develop instruments. However, the capability concept by Sen has been argued to be ambiguous concerning some elements of freedom, such as the burdens that people experience whilst achieving capabilities. Developing instruments with a comprehensive definition of capability might increase their sensitivity to a broader range of constructs. Our study operationalizes a framework based on the comprehensive “option freedom” concept into measurable constructs and presents an illustrative instrument.

For this, the Multi Instrument Comparison (MIC) database was used. Items from the MIC database were matched to themes from a framework that had been developed in an earlier qualitative study. Then, a measurement model was constructed with the selected items and model fit was assessed. Lastly, an illustrative instrument was created that shows how the selected constructs can be measured concisely.

A measurement model was constructed with 57 items and 11 factors. Data-driven explorative adjustments were made to improve model fit. Based on this model an instrument was developed with three scales (“Reflective Wellbeing”, “Affective Wellbeing” and “Perceived Access to Options”) totaling 15 items. This instrument showed adequate psychometric characteristics in terms of reliability and fit index values.

This study shows how the concept of option freedom can be operationalized for health-related wellbeing assessment. Furthermore, the analysis indicates that in the context of outcome measurement, information about both capabilities and functionings related to subjective wellbeing is required to assess the overall wellbeing of an individual. Further research is needed to validate the instrument.

## Introduction

The aim of health technology assessment (HTA) is to systematically examine the value of medical technologies. This is done by looking at their intended and unintended benefits and costs (Banta, 2003). Some HTA agencies require benefits to be measured in Quality Adjusted Life Years (QALY). QALYs are estimated with preference-adjusted scores of Health-Related Quality of Life (HRQoL) instruments. However, proponents of the capability approach argue that with their focus on health, the content of preference-adjusted HRQoL instruments is too narrow (Lorgelly et al., 2010). Indeed, it has been argued that these instruments are unable to assess the broader effects that medical technology can have on an individual’s life (Lorgelly et al., 2010). Instead, proponents of the capability approach argue that the benefits of medical technologies should be assessed in terms of their influence on the capabilities of individuals (Lorgelly et al., 2010). The concept of capability comes from the capability approach, which was initially developed by Amartya Sen (Sen, 1985). According to Sen, capabilities can be understood as the individuals’ freedom to achieve what they value. Sen argues that a comprehensive assessment of wellbeing should take these capabilities into account.

However, operationalizing concepts from the capability concept into measurable constructs is challenging. To illustrate, Ubels et al. (2022b) argue that some of the currently available capability instruments miss content related to the difficulties that individuals might experience when trying to fulfill their capabilities. This observation is supported by empirical analyses that have shown that some capability instruments are relatively insensitive to physical health problems (Engel et al., 2017, Davis et al., 2013, Hackert et al., 2017, Khan and Richardson, 2018). These problems might not block the achievement of capabilities but could significantly burden their realization, thus limiting the capabilities of individuals (Ubels et al., 2022b). To illustrate, an individual that has mobility problems might still be able to visit their friends, but it might take an extra-effort because of the need to call a taxi instead of taking a bicycle.

The relative insensitivity of these instruments to the difficulties that people might experience could be the result of them being based on a relatively narrow conceptualization of capability by Sen (1985). In his conceptualization of capability, Sen (1985) focused on the ability of individuals to achieve what they value. This conceptualization does not articulate the difficulties experienced by individuals while realizing their capabilities (Ubels et al., 2022b). A potential solution to this challenge is to develop an instrument that is based on a more comprehensive conceptualization of capability (Ubels et al., 2022b, Ubels et al., 2022a). Such a broader conceptualization was proposed by Robeyns (2017), who suggested that a capability is best understood as an “option freedom”. The concept of option freedom is developed by Pettit (2003), who argues that freedom is best understood as consisting of (1) **options**, which are the alternatives that an individual is in a position to realize, and (2) **access to options**, which refers to the ability of an individual to realize her/his options.

According to Pettit (2003), **options** should be understood as “*the alternatives that an individual is in a position to realize*”. Options are defined by several characteristics; for instance, for an individual, options have an **objective and subjective significance**. The objective significance is related to how an option affects an individual’s world. For instance, if two individuals each have a remote control, both have the option to press the bottom and change the television program. However, if the first individual has a remote control with empty batteries and the second individual has fully charged batteries, objectively, the second individual has more freedom than the first individual. An option’s **subjective** significance is related to the personal value of a particular option to an individual. Take as an example two individuals who decide between going to a football or a basketball match. Individual one is interested in sports; individual two is not. Then, it could be argued that individual one has more freedom, given that the choice between the different options matters more to her/him.

**Access to options** is understood as the ability of an individual to realize his/her options. This reflects the fact that access to options can be **blocked or burdened**. For instance, if an individual wants to ride her/his bike, but the bike is locked, and she/he lost the key, then the option to ride the bike is blocked. If, however, the key is not lost but left at home, then the access to the option of riding a bike is burdened, meaning that to realize this option, she/he needs to make an additional effort and go back home to get the key. Blocks and burdens of access can either be **objective** (such as the example of the key above) or **subjective**. An example of a subjective block could be that due to living in a patriarchal society, a woman believes that she is not able to drive a car, even though there are no objective blocks for her to do so.

The concept of option freedom was developed in the context of the philosophical debate on how freedom should be understood. The concept consequently needs further adaptation before it can be used as a basis for instrument development. This can be done by developing a theoretical framework that is based on the concept of option freedom, with themes that reflect the options that individuals value as well as the blocks and burdens that they experience.

Such a study was conducted by Ubels et al. (2022a). In this study, the concept of option freedom was used as an a-priori concept to interpret qualitative data using a best-fit framework synthesis (Carroll et al., 2013). Two a-priori concepts, ‘Options’ and ‘Access to Options’, were used to deductively analyze data from qualitative studies. Data that did not match the a-priori concepts were inductively analyzed to adjust the a-priori themes and, when necessary, develop new themes. Based on this analysis, a framework consisting of four themes was developed:

¬ **“Perceived Access to Options”** captures how individuals perceive their ability to realize valuable options from the range of options available to them. This theme essentially reflects various capabilities of individuals.
¬ **“Perceived Control”** is related to individuals’ perceived ability to influence their own lives.
¬ **“Option Wellbeing”** reflects different abstract options that people have access to that result in the experience of wellbeing. Examples of such options are “being satisfied with their health” due to having access to the option of being healthy (e.g., being able-bodied or not being in pain), “being happy” as a consequence of having access to options that result in happiness (e.g., hobbies), or “being satisfied with their social relationships” due to having access to options related to social relationships (e.g., having friends or family).
¬ **“Self-Realization”** represents the idea of having the experience of being able to do meaningful things in light of the options that individuals have access to, for example, “being independent” or “self-determination”.

The subjective experience of having access to options is then defined as a combination of the experiences described in the themes “Perceived Control”, “Self-Realization” and “Option Wellbeing”, which are essentially functionings. Consequently, this framework combines information about health-related capabilities and functionings to assess wellbeing.

The framework from Ubels et al. (2022a) has two advantages over Sen’s concept of capability, which forms the base of previously developed capability instruments. First, the framework acknowledges that two individuals with similar options cannot be considered to have an equal level of capability if one individual has more trouble accessing those options than the other. Second, the framework explicitly lists functionings that reflect key experiences for wellbeing assessments. This is in line with the observation by Clark (2005) that the capability approach should elaborate more on subjective experiences that could be important for an individual’s wellbeing.

The framework proposed by Ubels et al. (2022a) was designed in such a way that it can be operationalized as an instrument that can be used to assess wellbeing. It aims to operationalize the measurement of option-freedom in the context of health. Furthermore, the framework claims that there are benefits to assessing wellbeing in terms of both capabilities and experienced wellbeing, since each would, in theory, provide supplemental information about the overall wellbeing of an individual in the context of self-report instruments. This has however not been tested yet.

Therefore, the research presented in this manuscript has two aims. The first aim is to study whether the themes from Ubels et al. (2022a) can be operationalized as measurable constructs. By doing so, we also test the hypothesis whether capability and experienced wellbeing consist of different constructs that indeed provide supplemental information in the context of wellbeing assessment with self-report instruments. The second aim is to illustrate what kind of content might be included in a parsimonious capability instrument that operationalizes the concept of option freedom.

## Methods

In this study, a confirmatory factor analysis (CFA) was conducted (Kline, 2011). For this analysis, we used data from the Multi-Instrument Comparison (MIC) study (Richardson et al., 2012). The objective of the MIC study was to compare different general and disease-specific HRQoL and wellbeing instruments. The study followed a cross-sectional design and was conducted in six countries (Australia, Canada, Germany, Norway, the United Kingdom, and the USA) with 9,665 participants The MIC sample included groups of individuals affected by certain diseases (arthritis, asthma, cancer, depression, diabetes, hearing loss, and heart problems) as well as participants without disease. The MIC study team removed unreliable responses. Unreliable responses were identified based on inconsistencies in responses to items, as well as the time that it took for participants to complete the full MIC questionnaire (see the removal process explanation in Appendix Section 1). This resulted in a database of 8,022 evaluable responses. Further information about the MIC study can be found at Richardson et al. (2012) and the website of the MIC project (https://www.aqol.com.au/index.php/aqol-current).

Participants of the MIC study completed 11 different instruments in the main questionnaire, which in total consists of 227 questions or items. Items were structured as Likert - or Likert-like scales, with response options ranging from two to eleven. For our analysis, the responses to all items were recoded such that higher values mean that an individual is better off or is less limited.

### Item selection: Relevant items considered for use in the measurement of the four themes

Out of 227 items, we selected those which are relevant to the four themes “Perceived Access to Options”, “Perceived Control”, “Self-Realization” and “Option Wellbeing” (See Appendix Table A1 and Ubels et al. (2022a)). In this selection, the wording and content of the quotes supporting the identification of the four themes from Ubels et al. (2022a) were compared to the wording and content of the items included in the MIC questionnaires. Examples of quotes and selected items are presented in Appendix Table A1. Items with fewer than four response options were excluded from this selection to enhance measurement precision (Simms et al., 2019).

### Model development: Construct testing

We randomly split the MIC sample into two subsets. One subset of the data functioned as a training dataset for constructing the measurement model. The resulting model was validated on the second subset (hereafter *test dataset*).

Several measurement models were developed to test if the four themes could be operationalized as measurable constructs. These models were tested using a robust maximum likelihood (MLR) estimator and developed in two stages:

1. A *theory-driven confirmatory stage:* A measurement model was developed and tested to see if the hypothesized themes can be operationalized as constructs in an instrument. Further theoretically guided adjustments were made to see if the model fit of the measurement model could be improved.
2. A *data-driven explorative stage:* Further data-driven adjustments to the measurement model were guided by studying model misfit. First, the patterns of residual correlations of items with values higher than 0.1 and modification indices were studied to identify (local) misfit (Kline, 2011). Second, when misfit was identified, the content of the items and their layout in the MIC questionnaire were studied. Third, when the content of items showed similarities or the layout of the MIC questionnaire could have caused additional correlations amongst sets of items, specific orthogonal factors were created. In the case of two items showing misfit and sharing content, the errors of the two items were correlated. Lastly, a model was developed with orthogonal factors to account for common method variance caused by similarities in the number of response options (Podsakoff et al., 2012).

The theoretical and empirical adjusting stages resulted in the selection and development of a final measurement model. The robustness of the model fit was tested in two stages:

1. The final model was validated with the test dataset.
2. The final model was estimated with an estimator suitable for analyzing categorical data to test the robustness of the model fit test results. Items with up to seven response options were treated as categorical variables. The final measurement model was then estimated with a Diagonally Weighted Least Squares (DWLS) estimator and polychoric correlations for items with up to seven response options.

Model fit was examined with scaled versions of the χ^2^, Comparative Fit Index (CFI), Tucker Lewis Index (TLI), Root Mean Squared Error of Approximation (RMSEA), and Standardized Root Mean Residual (SRMR) fit indices. Values higher than 0.9 for the CFI and TLI fit indices indicated acceptable fit, with values closer to 0.95 being preferable (Hu and Bentler, 1999). A RMSEA value lower than 0.6 and an SRMR lower than 0.8 were also used to indicate acceptable fit (Hu and Bentler, 1999). Nested model comparisons were made according to the guidelines of Chen (2007), who argues that differences in CFI, SRMR, and RMSEA values larger than 0.01, 0.015, and 0.015, respectively, indicate a substantial improvement in model fit. Smaller differences in these values represent a negligible difference in model fit, in which case the more parsimonious measurement model was considered superior.

Missing data were handled with a Full Information Maximum Likelihood (FIML) estimator for the models estimated with a MLR estimator and pairwise deletion for the model estimated with a DWLS estimator.

The software used for this analysis was the Lavaan package version 0.6-10 (Rosseel, 2012) in R version 3.5.2 (R Core Team, 2013). Additional information on the development of the final measurement model is provided in Appendix Section 1.

### Item selection for an illustrative instrument for the assessment of capability wellbeing

An illustrative instrument was created that was based on the items included in the final measurement model in three steps:

1. First, the inter-factor correlations were studied. Correlations higher than 0.9 indicate that two constructs are closely associated with each other from a measurement perspective. To develop a parsimonious instrument, Le et al. (2010) suggest retaining only one of such correlated constructs, since measuring one construct sufficiently captures information about the other(s). Accordingly, we retained one construct for the instrument when such highly correlated factors were identified.
2. Second, items that cross-load on multiple factors were removed.
3. Third, one item was retained from groups of items that covered similar content. These items were selected on a case-by-case basis. Special attention was paid to the ceiling and floor effects of individual items, the variability of the responses over the different response options, and the content of the items. We also evaluated the item-total correlations, which showed negligible differences between items and, therefore, were not used in the item selection procedure. A detailed explanation of the selection process is provided in Appendix Section 2.

This instrument was developed with the full dataset. The aforementioned fit indices (i.e., χ^2^, CFI, TLI, RMSEA, and SRMR) were used to assess the measurement model fit of the developed instrument. Cronbach’s alpha values were also computed for the instrument’s scales (Cronbach, 1951).

## Results

Out of the 8,022 observations, 1,191 contained missing values. Most of the missing values (*n* = 1,177) corresponded to the Norwegian participants’ responses to the ICECAP-A and AQoL–4D instruments. Norwegian translations of these instruments were unavailable at the time of data collection. The analysis of the data suggests that the probability of being missing is the same within the group of observations from the same country, and therefore, this data was missing at random. Table 1 shows the characteristics of the included MIC survey participants.

**Table 1.** Demographics of the MIC study sample.

|  |  | **N (%)** |  |
| --- | --- | --- | --- |
| **Total** |  | **8,022** | **(100%)** |
| Age |  |  |  |
|  | 18-24 | 513 | (6.4%) |
|  | 25-34 | 944 | (11.8%) |
|  | 35-44 | 1,137 | (14.2%) |
|  | 45-54 | 1,689 | (21.1%) |
|  | 55-64 | 2,008 | (25.0%) |
|  | 65+ | 1,731 | (21.6%) |
| Gender |  |  |  |
|  | Male | 3,848 | (48.0%) |
|  | Female | 4,174 | (52.0%) |
| Education |  |  |  |
|  | High school | 2,522 | (31.4%) |
|  | Some post-secondary, post- secondary certificate or diploma | 3,241 | (40.4%) |
|  | University degree and higher | 2,259 | (28.2%) |
| Disease |  |  |  |
|  | Healthy public | 1,760 | (21.9%) |
|  | Arthritis | 929 | (11.6%) |
|  | Asthma | 856 | (10.7%) |
|  | Cancer | 772 | (9.6%) |
|  | Depression | 917 | (11.4%) |
|  | Diabetes | 924 | (11.5%) |
|  | Hearing problems | 832 | (10.4%) |
|  | Heart problems | 943 | (11.8%) |
|  | Stroke | 23 | (0.3%) |
|  | Chronic obstructive pulmonary disease | 66 | (0.8%) |
| Country |  |  |  |
|  | Australia | 1,430 | (17.8%) |
|  | Canada | 1,330 | (16.6%) |
|  | Germany | 1,269 | (15.8%) |
|  | Norway | 1,177 | (14.7%) |
|  | United Kingdom | 1,356 | (16.9%) |
|  | United States of America | 1,460 | (18.2%) |

### Item selection: Assessing the relevance of items used to measure the four themes

We selected 56 items from the MIC database based on the similarities in their content with quotes from the theoretical framework developed in Ubels et al. (2022a). Details of the selected items and the respective quotes are provided in Appendix Table A1. Relevant items for measuring the four themes were identified in seven out of the eleven instruments included in the MIC database (see Appendix Section 1 and Appendix Table A1).

Out of the 56 items, 26 were linked to the theme “Option Wellbeing”, 20 to the theme “Perceived Access to Options”, 6 to the theme “Self-Realization”, and 4 to the theme “Perceived Control”. For the subthemes “Access due to Social Wellbeing”, “Access due to Activity Wellbeing” and “Access due to Finances”, from the theme “Perceived Access to Options”, no items were identified (See Appendix Table A1). Similarly, no items were identified for the “Having Dignity” subtheme from the theme “Self-Realization”.

### Model development: Model fit of four constructs

#### Stage 1. Theory-driven model development

The first measurement model consisted of four oblique factors representing each of the four themes. Model 1 in Table 2 displays the associated fit indices. The RMSEA, SRMR, CFI, and TLI indices of this model indicated inadequate model fit. For this reason, several theoretical re-specifications were made. These theoretical specifications concerned (1) two items that require participants to evaluate their own health, which were cross-loaded on the “Perceived Access to Options” factor, and (2) six items that had a testlet format (a bundle of items that follow one introduction, i.e. “Did your health influence the following activities” followed by three Likert-scale items describing different activities), for which orthogonal specific factors were developed (details provided in Appendix Section 1). These theoretical re-specifications improved the model fit (Table 2, Model 2); however, the RMSEA, SRMR, CFI, and TLI indices still indicated misfit. To improve the model fit further, we conducted a data-driven explorative analysis.

**Table 2.** Fit statistics of tested models.

| | Dataset | $\chi^2$ | df | RMSEA**<br>(90% CI) | SRMR<br>*** | CFI† | TLI† |
| --- | --- | --- | --- | --- | --- | --- | --- |
| **Model 1:** A-priori model | training dataset | 43,477.9 | 1,478 | 0.095<br>(0.095 – 0.096) | 0.100 | 0.727 | 0.711 |
| **Model 2:** After theoretical re-specifications | training dataset | 30,574.0 | 1,462 | 0.079<br>(0.079 – 0.080) | 0.096 | 0.814 | 0.804 |
| **Model 3:** After post-hoc adjustments with method factors for item length | training dataset | 9,501.9 | 1,368 | 0.043<br>(0.042 – 0.044) | 0.045 | 0.947 | 0.943 |
| **Model 4:** Final model without method factors for response option length | training dataset | 10,713.8 | 1,379 | 0.046<br>(0.045 – 0.047) | 0.045 | 0.939 | 0.935 |
| **Model 5:** Final model MLR* | test dataset | 10,819.4 | 1,379 | 0.046<br>(0.045 – 0.047) | 0.044 | 0.940 | 0.936 |
| **Model 6:** Final model DWLS* | test dataset | 14,672.3 | 1,379 | 0.053<br>(0.052 – 0.054) | 0.043 | 0.964 | 0.961 |

Comparative Fit Index (CFI), Tucker Lewis Index (TLI), Root Mean Squared Error of Approximation (RMSEA), Standardized Root Mean Residual (SRMR), degrees of freedom (df)
\* Models 1 to 5 were estimated with a Robust Maximum Likelihood estimator (MLR), model 6 with a Diagonally Weighted Least Squares (DWLS) estimator.
\*\* Values lower than 0.6 indicate of acceptable fit.
\*\*\* Values lower than 0.8 indicate of acceptable fit.
† Values higher than 0.9 indicate acceptable fit.

#### Stage 2. Data-driven model development

In the data-driven explorative analysis, four sources of misfit were identified.

1. Items having similarities in content beyond what was accounted for by the existing factors in the model (e.g. a group of items specifically asking about the need for help or support, the covariance which was not captured by the more general “Perceived Access to Options” factor).
2. The design of the instruments themselves, since some of the items were presented in a testlet format, which could result in additional covariance.
3. Miss-specified factor loadings, with some of the items cross-loading on multiple factors and others more appropriately loading on other factors than hypothesized.
4. Common method variance related to response option length of items in some questionnaires.

Two different measurement models were developed to account for these four sources of misfit. One model (Model 4 in Table 2) accounted for the first three sources of misfit, while the other accounted for all four (Model 3 in Table 2). The chi-square test suggests that Model 3 fits better than Model 4 (Δ df = 11, Δ χ^2^ = 1,211.9, p < 0.001). Nevertheless, given that the chi-square test is sensitive to large sample sizes, it is unhelpful to inform the current model comparison (Chen, 2007). The CFI, the SRMR, and the RMSEA showed a comparable fit between both models, with differences being smaller than 0.01, 0.015, and 0.015, respectively. Given the sensitivity of the chi-square test, the negligible differences in model fit shown by the other fit indices, and the fact that Model 4 is more parsimonious than Model 5, we retained Model 4 as the final measurement model.

A substantial difference between Model 1 and Model 4 was how the subjective experience of capabilities was structured. In the first a priori measurement model (Model 1), items from the MIC database were loaded on four factors that represented each of the four themes (see Appendix Table A1). The “Self-Realization” construct included items related to individuals’ experience of living a worthwhile life (e.g., code ONSj, Table A1) or having the idea that life is close to an ideal (e.g., code SWLS_a, Table A1). In Model 4, these items moved to the newly developed construct “Reflective Wellbeing”. Similarly, items that belong to Option Wellbeing in Model 1 and were related to satisfaction with different elements in life, were included in Model 4 in the “Reflective Wellbeing” construct (examples of such items are: satisfaction related to general life with codes ONSi and PWI_a, satisfaction with health with the code PWI_c, Table A1) or satisfaction with social relationships with the code PWI_e, Table A1). Additionally, the items of the “Option Wellbeing” factor that belong to the subtheme “Emotional Wellbeing”, in Model 4 were loaded on their own factor, which was relabeled to “Affective Wellbeing”. These items covered various emotional experiences, such as feelings of sadness (e.g., code aqol5, Table A1), anxiety (e.g., code sf24, Table A1), or pleasure (e.g., code aqol25, Table A1). A detailed explanation in how Model 1 and Model 4 differ from each other (in terms of added orthogonal factors and correlated erros) can be found in the Appendix.

The robustness of the fit index values of measurement Model 4 was tested with the test dataset, which resulted in Model 5 (see Table 2). As mentioned above, a comparison of the fit index values of Model 4 and Model 5 showed that the differences in model fit are negligible. A further robustness test was conducted by estimating Model 4 using a DWLS estimator and treating items with up to seven response options as categorical, which resulted in Model 6. The similarities in fit indices after estimating Model 6 (see Table 2) indicated that the estimations related to Model 4 and Model 5 were robust.

The standardized factor loadings of the items of measurement Model 5 are presented in Appendix Table A2 and the item error correlations can be found in Appendix Table A3. The inter-factor correlations of measurement Model 5 can be found in Appendix Table A4. The results suggest that the correlation between the factors “Perceived Control” and “Affective Wellbeing” is 0.911. The other standardized factor correlations were moderate, ranging from 0.395 to 0.756.

### Instrument development

From the 56 items of the measurement models presented above, 15 were selected to form an illustrative instrument that assesses capability wellbeing in terms of three constructs: “Perceived Access to Options”, “Reflective Wellbeing”, and “Affective Wellbeing”. The decision to exclude the “Perceived Control” was based on inter-factor correlations. The analysis showed a high correlation (> 0.9) between the “Perceived Control” and “Affective Wellbeing” constructs; therefore, measuring one of the constructs provides sufficient information about the other construct (Le et al., 2010).

The remaining three scales assess wellbeing in terms of health-related capabilities with the “Perceived Access to Options” scale and experienced wellbeing in terms of the **“**Affective Wellbeing” scale and the “Reflective Wellbeing” scale. The three scales consist of 15 items: “Perceived Access to Options” (Cronbach’s alpha: 0.89, 5 items), “Affective Wellbeing (Cronbach’s alpha: 0.83, 4 items), and “Reflective Wellbeing” (Cronbach’s alpha: 0.89, 6 items). The fit of the measurement model of the instrument was adequate (χ^2^: 1,756.8, df: 87, CFI: 0.970, TLI: 0.963, RMSEA: 0.055, SRMR: 0.036). The three scales and their corresponding items are listed in Appendix Section 3.

The 15 items were selected in such a way that they minimize floor and ceiling effects. We estimated the floor and ceiling effects based on the MIC study sample. For the “Perceived Access to Options”, “Reflective Wellbeing” and “Affective Wellbeing” scales, the floor effects were 0.01%, 0.04%, and 0.25%, and the ceiling effects were 15.76%, 0.96%, and 2.68% respectively. The proportion of participants per response option and item is provided in Appendix Table A5.

The illustrative instrument developed in this study is titled the Wellbeing Related option-Freedom (WeRFree) instrument. Figure 1 is a graphic representation of the measurement model of the WeRFree instrument. Further results of the CFA conducted with the measurement model of the WeRFree (standardized factor, standardized intercepts, and standardized variances of the items on their respective scales) can be found in Appendix Table 6. Additionally, Appendix Table 7 presents item-total correlations per scale.

**Figure 1.**
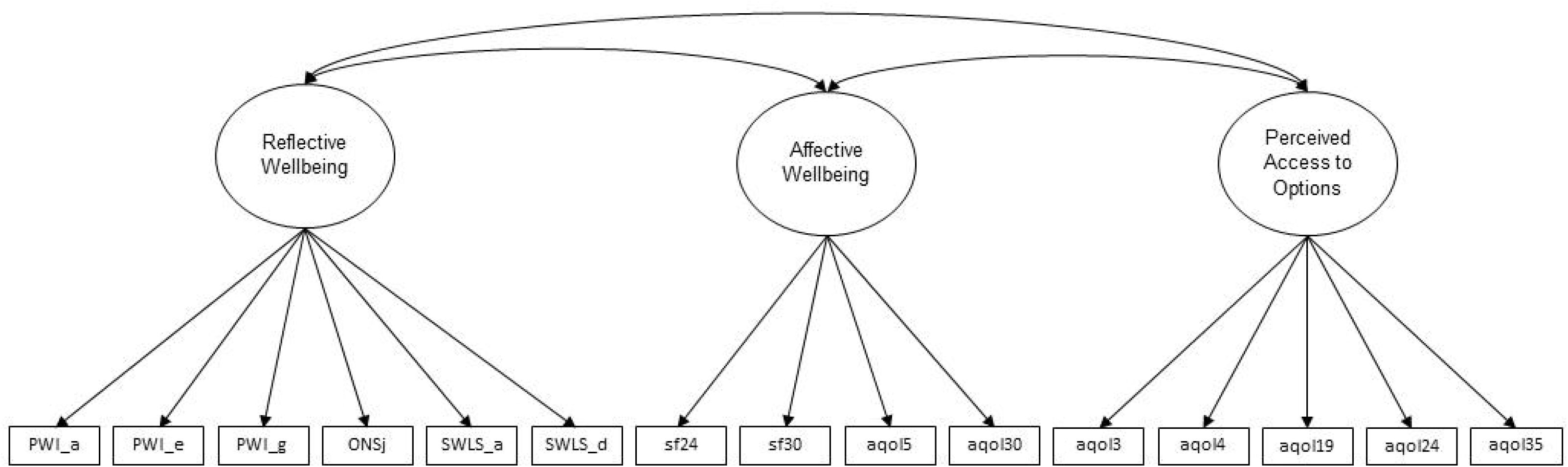
Graphical presentation of the WeRFree measurement model Note: The item linked to the codes can be found in Appendix Table 2

## Discussion

The study described in this article had two aims: studying whether the themes from the theoretical framework developed by Ubels et al. (2022a) could be operationalized as measurable constructs and presenting what a parsimonious instrument that is based on the concept of option freedom might look like.

Regarding the first aim, the first two versions of the theoretical measurement model that included all items were based on the theoretical framework from Ubels et al. (2022a). The poor fit of the resulting models indicated that the internal structure of the constructs was different than hypothesized in the theoretical framework (Ubels et al., 2022a). Consequently, explorative data-driven adjustments were performed to assess the structure of the larger measurement model and study if model fit could be improved. The resulting model that showed the best fit differed from the original qualitative framework in how the subjective experience of capabilities was structured. In their framework, Ubels et al. (2022a) suggested that two themes capture the subjective experience of capabilities: “Option Wellbeing” (the happiness and satisfaction that people experience when their options are fulfilled to an adequate level) and “Self-Realization” (the experience of living a meaningful life). However, explorative data analysis suggested that a better model fit could be achieved by structuring experienced wellbeing as reflective and affective wellbeing. This meant that items that reflect life satisfaction were combined with items that reflect living a meaningful life, instead of being related to a construct that reflects various emotional aspects of wellbeing. “Reflective Wellbeing” refers to cognitive appraisals about an individual’s wellbeing, such as having a sense of life satisfaction and meaning. “Affective Wellbeing” represents the emotional aspects of wellbeing, such as happiness and sadness.

Structuring the subjective experience of wellbeing as consisting of a cognitive (“Reflective Wellbeing”) and an emotional (“Affective Wellbeing”) construct is in line with insights from the literature (Diener, 1984). The psychological literature illustrates that affective wellbeing could further be subdivided into positive and negative affect (Lucas et al., 1996, Diener, 1984, Busseri and Sadava, 2011). However, how these constructs are structured amongst themselves is still under debate (Busseri and Sadava, 2011). In our study, the bifactor-like approach used in the CFA can be considered to be an adequate method of modeling subjective wellbeing (Jovanović, 2015).

Regarding the capability construct, the best-fitting larger measurement model was consistent with the original theoretical framework. In both, the measurement of capabilities was operationalized with the construct “Perceived Access to Options”. This construct assesses how individuals perceive their ability to access options and captures the difficulties that people experience while achieving their capabilities (e.g., pain). These difficulties are reflected in HRQoL questionnaires. This idea is consistent with Cookson (2005), who observed that some of the items in the EQ-5D (an HRQoL instrument commonly used to calculate QALYs) might reflect health-related capabilities. Taking the item “usual activities” from the EQ-5D as an example, Cookson argues that individuals can only respond to this item by reflecting on the effect of health on non-health-related functionings. Thus, in this context, questions around health can be used to understand individuals’ health-related capabilities.

The factor “Perceived Access to Options” showed a low-moderate correlation with “Reflective Wellbeing” (0.406) and “Affective Wellbeing” (0.530). This suggested that the health-related capabilities and the subjective wellbeing of individuals are two distinct elements of wellbeing. Therefore, an instrument solely measuring capability conceptualized as option freedom might be less sensitive to functionings related to the subjective wellbeing of individuals. This is in line with the observation by Clark (2005) that wellbeing assessment might be improved by combining information about capabilities and the subjective wellbeing derived from having those capabilities. Consequently, when assessing wellbeing, the measurement of both health-related capabilities and subjective wellbeing constructs would result in a more comprehensive assessment of wellbeing than the assessment of each of them separately.

Regarding the second aim of this study, the WeRFree instrument, illustrates how wellbeing can be assessed with a parsimonious instrument that is based on the concept of option-freedom. Additionally, it has content that reflects individuals’ experienced wellbeing. To achieve this parsimony, we decided to include the “Affective Wellbeing” construct and exclude the construct “Perceived Control” in the instrument. The decision to favor the “Affective Wellbeing” construct was based on theory. In the article by Ubels et al. (2022a), it was hypothesized that “Perceived Control” is a key construct that influences how individuals experience their capabilities, which is represented by the outcome variables “Reflective Wellbeing” and “Affective Wellbeing”. As such, “Perceived Control” is not necessarily an outcome variable, which meant that instead the “Affective Wellbeing” construct was included in the instrument.

Compared to capability instruments based on Sen’s conceptualization of capability, the WeRFree instrument is potentially more sensitive to the health-related burdens that people experience than existing capability instruments. Furthermore, the WeRFree instruments allows differentiating between elements of wellbeing linked to capabilities and the subjective experience of wellbeing, which could be seen as a functioning. Sen’s conceptualization of capability, with its focus on freedom, might impede researchers from identifying other relevant aspects of wellbeing (Ubels et al., 2022b).

Regarding conventional health economic instruments, studies suggest that some HRQoL instruments are relatively insensitive to the psychosocial effect of health technologies due to their focus on (physical) health (Khan and Richardson, 2018, Richardson et al., 2015). Compared to these instruments, the WeRFree instrument should more comprehensively assess the wellbeing of individuals, due to its reflective and affective wellbeing constructs. (Engel et al., 2017, Davis et al., 2013, Hackert et al., 2017, Khan and Richardson, 2018).

## Limitations

The MIC study database, used for our item selection, covered most of the themes and subthemes proposed by Ubels et al. (2022a). However, it did not contain items that could be linked to the subthemes “Access due to Social Wellbeing”, “Access due to Activity Wellbeing”, “Access due to Finances”, and “Having Dignity”. In the context of the WeRFree instrument, further research is necessary to examine if items related to these subthemes should be measured as independent constructs or can be incorporated in the existing scales of the WeRFree instrument.

Another limitation is related to the explorative nature of the current research. The values of the models’ fit indices estimated with the training dataset should not be interpreted as confirming or rejecting hypotheses concerning the ability of the measurement model to predict the covariance structure of the data (Wagenmakers et al., 2012). Additionally, the model’s fit indices estimated with the test dataset should be interpreted with caution since both the training and test dataset share the measurement errors that are caused by the research design. This could lead to an overestimation of the models’ fit in both datasets. Further research with external datasets should be conducted to confirm the model’s fit.

In addition, a capability instrument’s content essentially functions as a capability list (Robeyns, 2005). As a capability list, further procedural steps (such as a public defense of the content) need to be followed before the instrument can effectively be used to inform policy-making (Robeyns, 2005). Consequently, the WeRFree instrument should be considered as an illustrative instrument that will require further development and validation before it can be used in policy-making.

## Conclusion

In light of the importance of accurately and comprehensively assessing the value of new medical interventions, the content of a capability instrument used with this aim should be broad enough to measure new technologies’ effects accurately. In this context, researchers interested in the assessment of wellbeing should consider using a comprehensive concept to operationalize capability for wellbeing assessment and combine this information with subjective wellbeing, since both yield unique information that should be used for policy making in specific contexts, such as the assessment of the effects of health technologies.

The WeRFree instrument, developed in this study, illustrates what kind of content might be included in such an instrument. Further research is however needed to validate and explore the properties of the WeRFree instrument.

## Data Availability

All data produced in the present study are available upon reasonable request to the study team of the Multi Instrument Comparison (MIC) project, which can be contacted at: https://www.aqol.com.au/index.php/mic-data

## Declarations

No funding was received to assist with the preparation of this manuscript.

