## Appendix for "The value of freedom: the introduction of option freedom in health-related capability wellbeing measurement"

### Appendix Section 1: Further information about results and model development

#### Identification of unreliable responses

The study team that developed the MIC dataset removed unreliable responses. Unreliable responses were removed in eight rounds of edits (Richardson et al. 2012). In round (1), responses were removed from participants who completed the full MIC questionnaire in less than 20 minutes. Responses by participants who took between 20 and 25 minutes to complete the survey were further scrutinized. For round (2), the mobility item of the EQ-5D-5L was presented two times in different places. Responses were removed when the difference between the responses to the items was more than two points on the Likert scale. When the difference was only one point, the responses of the participant were further put under further scrutiny. For round (3), responses to two items were compared: the first item of the 36-Item Short Form Health Survey V2 (SF-36, Brazier, Roberts, and Deverill (2002) and another item not belonging to an instrument. These items asked participants to self-assess their general health. Responses that differed by more than one point on the Likert scale were removed from the database. Responses that differed by one point were noted for further inspection. In round (4), responses to the first item of the SF-36 and the Quality of Well Being Self-Administered instrument (QWB-SA), developed by Seiber et al. (2008), were compared since both cover the self-assessment of general health. Similar to round (3), responses that differed by more than one point on the Likert scale were removed and responses that differed by one point were noted. Round (5) was again a comparison between two items from the QWB-SA and the self-assessment of the general health item mentioned in round (3). Responses that differed by more than one point were deleted and responses that differed by one point were noted for further inspection. In round (6), responses to item 22 of the AQoL-8D and the fourth item of the EQ-5D-5L were compared to each other, which both cover the experience of pain. Responses were removed from the database when they differed by two or more points on the Likert scale. For rounds (7) and round (8), the items that were noted for further inspection in rounds (1), (2), (3), (4), (5), and (6) were scrutinized. Inconsistencies in these rounds were summed (i.e. a one point Likert-scale difference in rounds (1) and (2) resulted in a score of two). In round (7), responses were deleted from the database when two inconsistencies were identified and the participant completed the full MIC survey in less than 25 minutes. In round number (8), responses were removed when three or more inconsistencies were identified.

#### Item selection

A total of 57 items from seven questionnaires were selected to develop the measurement model. Three of these questionnaires were HRQoL instruments: (1) 36-Item Short Form Health Survey V2 (SF-36, Brazier, Roberts, and Deverill (2002);, and (2) the Assessment of Quality of Life instrument (AQoL) with four dimensions (AQoL-4D, (Hawthorne, Richardson, and Osborne 1999), and (3) the AQoL with eight dimensions (AQoL-8D, (Richardson et al. 2009). Three were subjective wellbeing instruments: (1) Satisfaction With Life Scale (SWLS, (Diener et al. 1985), (2) the Integrated Household Survey from the Office of National Statistics (ONS, (Dolan and Metcalfe 2012), and (3) the Personal Wellbeing Index (PWI, (Cummins et al. 2003). Additionally, items from the ICEpop CAPability measure for Adults (ICECAP-A, (Al-Janabi, N Flynn, and Coast 2012) were also used. Appendix Table 1 presents the themes and associated items from the MIC database.

#### Theoretical adjustments to the measurement model

Several theoretical specifications were made to study if model fit could be improved. Items SF1 and PWI_c both cover whether individuals experience satisfaction with their health or consider their own health to be excellent. Because the factor “Perceived Access to Options” contains items that reflect the influence of physical or mental health on capabilities, it was hypothesized that these two items would cross-load on this factor.

Lastly, based on the layout and the wording of the MIC questionnaire, it was hypothesized that the items sf13, sf14, sf15, and sf16 shared covariance and the items sf17, sf18, and sf19 shared covariance. For this reason, a separate orthogonal factor was created which predicted a part of the variance of sf13, sf14, sf15, and sf16, and a separate orthogonal factor was created that predicted the covariance of items sf17, sf18, and sf19. The fit index values of this model can be found in Table 2 in the main manuscript as model 2.

#### **Data-driven** adjustments to the measurement model

The fit indices indicated that the model could be improved further. It was hypothesized that the model did not adequately account for all the covariance in the dataset due to similarities in wording amongst some of the items. Another possibility was that the theoretical model itself was misspecified. To study the sources of misfit and see if the theoretical model should be adapted, it was decided to study the residual correlations amongst the items to identify the sources of misfit and check modification indices.

First, the structure of subjective wellbeing was changed. Originally, subjective wellbeing was conceptualized as consisting of a factor that covers both satisfaction with life and emotional aspects of wellbeing, and a factor that covers the ability to do things that have meaning. Combining the items related to life satisfaction and a perceived sense of meaning into a new factor called “Reflective Wellbeing” improved model fit, with the factor with left over items associated with the experience of a variety of emotions being renamed into “Affective Wellbeing”.

Second, some items were made to cross-load over multiple factors. Several items of the factor “Perceived Access to Options” were made to cross-load on “Affective Wellbeing”, with the reason that these items were measuring limitations in access due to emotional problems (sf17, sf18, and sf19). It was hypothesized that these items would load on both the factors “Perceived Access to Options” and “Affective Wellbeing”.

Other items were also allowed to cross-load. The items aqol23 and aqol27, which load on the factor “Affective Wellbeing” were also allowed to load on the factor “Reflective Wellbeing”. The items sf29 and aqol26 were also allowed to cross-load on the factor “Perceived Access to Options”. The items aqol5_4D and aqol10 were respecified to also load on the factor “Affective Wellbeing”. The item aqol5_4D was deleted from the factor Reflective Wellbeing. One item, ONSk, was completely deleted. This item asks about the experience of happiness. However, the item loaded on the “Reflective Wellbeing” factor (standardized loading of 0.660) and only weakly on the affective factor (standardized loading of 0.185). Based on the layout of the questionnaire, it might be possible that answering the other two ONS items (ONSi and ONSj) influenced the response on the ONSk item, which are both items that also show high loadings on the “Reflective Wellbeing” factor. As such, it seems that the content of the item and what participants are answering are not in line with each other. As a consequence, we decided to delete the item ONSk.

A notable diversion of the a-priori hypothesized measurement structure are three items from the ICECAP-A questionnaire, developed to assess capabilities (for the included items, see Appendix Table 1); items ic03 (about being independent) and ic04 (about achievement and progress) initially loaded on the factor “Self-Realization”. Item ic05 loaded on the factor “Option Wellbeing”. However, the residual correlations showed that model fit could be improved if item ic03 loaded on both the factors “Perceived Access to Options” and “Perceived Control”. Similarly, item ic05 was allowed to cross-load on the “Perceived Control” factor and the “Affective Wellbeing” Factor. Lastly, item ic04 was respecified to load solely on the factor “Perceived Control”. The fact that these three items load on the “Perceived Control” factor might be due to the wording of questions, such as “I am able to” and “I can”. Similarly worded questions have been used in the field of psychology to measure constructs such as self-efficacy (Frei et al. 2009), which represents an individual’s perception of being able to do things (Bandura 2001). Additionally, item ic03 was allowed to cross-load on the “Perceived Access to Options” factor. Residual correlations indicated that this item showed unexplained covariance with multiple items in this factor. The perceived ability to access options seems to influence the perceived ability to be independent.

Third, seven specific factors were created to account for covariances due to similarities in content and the layout of the MIC questionnaire: (1) a factor that accounts for covariance regarding social aspects, (2) a factor that accounts for covariance between items that inform about the need for help or support, (3) a factor which accounts for covariance due to items inquiring about happiness, (4) a factor which accounts for covariance between items asking about anxiety, (5) a factor that accounts for the covariance of the physical limitations testlet from the SF-36, (6) a factor that accounts for the covariance of the emotional limitations testlet from the SF-36 and (7) a factor which accounts for similarities between the items of the SF-36 which assess negative emotions, such as feeling down or depressed. Because of the similarities in wording and the design of the instrument, the decision was made to let physical limitations and emotional limitations testlets of the SF-36 correlate with each other. The other factors were orthogonal.

Some errors of items were allowed to correlate, as was mentioned in the explanation about the general steps that were taken to explore the data in the main body of this study. Besides the items PWI_c and sf1, of which the errors were correlated because they both cover general health perceptions, also the errors of the items aqol24 and sf22 were correlated with each other. Both these items inquire whether pain influences normal activities or normal work. The errors of items sf20 and sf32 were also correlated with each other. These items were very similar since both ask how health and emotional problems interfered with social activities. Lastly, the errors of the items aqol2_4D and aqol30 were correlated with each other, since they both cover the need for help with household tasks. The correlation values of these error terms can be found in Appendix Table 3 (measurement model 5, estimated with the test dataset).

Lastly, a model was developed with method factors that accounted for similarities in the length of response options of items. This measurement model could not successfully be estimated, however, a simplified model that accounted for some common method variance was successfully constructed. This resulted in a measurement model with three additional specific orthogonal factors. One of these factors accounted for covariance between items of the SWLS questionnaire, another factor accounted for covariance between items from the PWI questionnaire and the last specific factor accounted for covariance between items from the ICECAP-A. The fit indices associated with this measurement model are presented as model 3 in Table 2 in the main manuscript. However, adding these three factors to account for similarities in item length resulted in a complex model. For the sake of parsimony, we decided to test model fit by removing the SWLS, the PWI, and the ICECAP-A specific factors. This resulted in measurement model 4 (Fit index values presented as model 4 in Table 2 in the main manuscript).

### Appendix Section 2: Further information about instrument development

From the pool of 56 items, 38 items remained after removing the “Perceived Control” constructs and deleting the items that cross-loaded on the factors “Reflective Wellbeing”, “Affective Wellbeing” and “Perceived Access to Options”. From these 38 items, a selection was made for which items to retain. The aim of this selection was to retain one item from a group of items that covered similar content. For example, only one item would be retained from three items that ask about the experience of pain.

As a start, one item was selected from a group of items that cover the ability of individuals to take care of themselves: AQOL1_4D, AQOL3_4D, and AQOL19. From this group of items, the decision was made to delete the items with the code AQOL1_4D and AQOL3_4D, because for these items around 85% of the participants chose the optimal response option, while in AQOL19 only around 66% chose the optimal response option. Given that the aim was to create a parsimonious instrument, it was decided to drop these items, since they did not give a lot of information about a large part of the population.

The items which cover various experiences of happiness were also scrutinized: ic05, sf30, aqol20, and aqol25. This group reflects the “positive” experience of affect, which meant that for the development of an instrument extra attention was paid to ceiling effects. Of these four items, item ic05 was removed first, given its high ceiling effect compared to the other items (0.34 vs 0.11, 0.08, and 0.11 respectively). The three remaining items showed a high proportion of responses clustering around their respective two optimal response options (sf30: a proportion of 0.69 vs. aqol20: proportion of 0.83 and aqol25: proportion of 0.80). Still, item sf30 showed the lowest proportion of clustering, thus it was decided to keep the item for the instrument.

Another group of items was related to the experience of anxiety: aqol11_4D, ONSl, and sf24. First, the item ONSl was removed from this group of items, because it asks about the experience of anxiety for only “yesterday”. This was not in line with the other items in the Affective Wellbeing scale, which did not specify a recall time, so the item was removed for the sake of consistency. Item sf24 was kept. Again, this was due to a high proportion of the respondents clustering their responses to the two “optimal” response levels, with sf24 showing the lowest proportion (sf24: proportion of 0.71, aqol11_4D: proportion of 0.80)

The items sf24, sf25, and sf28 also formed a group of items that shared variance, since they were part of a subscale of the SF-36 that assesses emotional wellbeing. As mentioned in the last paragraph, sf24 was kept, given that it reflects the anxiety that individuals feel. In the decision between sf25 and sf28 special attention was paid to floor effects, given that these items reflect “negative” experiences of affect. Following this, it was decided to keep sf25, given that it has a lower floor effect (sf25: 2%, sf28: 4%).

The items aqol5_4D, aqol10, aqol23, aqol34, and PWI_e were all linked to relationships. From these items, it was decided to keep PWI_e for two reasons. One, the responses were more evenly spread compared to the other items. Two, the items aqol5_4D, aqol10, aqol23, and aqol34 each have “inconsistent” response options. For example, the content of item aqol5_4d covers “having plenty of friends” and “feeling lonely”, both of which could be seen as different constructs.

The last cluster of items was sf13, sf14, sf15, sf16, and sf22. These items reflect physical health problems that interfere with accessing a variety of options, such as being limited in work or activities. The items sf13, sf14, sf15, and sf16 were constructed as a testlet and were not included in the instrument, since including such a number of items would conflict with building a parsimonious instrument. Therefore, it was decided to initially keep sf22.

Besides larger clusters of items that were linked to specific factors, also a couple of items shared variance, which was modeled by correlating their error terms. Also from these items, only one item was kept. In the following paragraph, it is explained how from these pairs items were selected for inclusion in the WeRFree instrument.

Items aqol24 and sf22 assess the impact of pain on usual activities and normal work respectively. From these two items, it was decided to drop sf22. The factor loadings of both the items were similar (standardized factor loading of aqol24 is 0.776, loading of sf22 is 0.784). However, responses were more evenly spread over the different answer options of aqol24, indicating that item aqol24 was slightly better at differentiating between different levels of the effect of pain in different groups of participants. Items aqol2_4D and aqol30 assess the need for help when doing household tasks. From these two, it was decided to drop aqol2_4D, because almost 70% of the participants chose its optimal response option (compared to 47% for item aqol30).

Appendix Table A1. Original themes, quotes related to those themes, items from the database with their code and the original instrument to which these items belong.

| Theme | Subtheme | Quotes | ITEMS mic database | MIC code | Instrument |
| --- | --- | --- | --- | --- | --- |
| Perceived access to options | Items directly linked to theme |  | During the past 4 weeks, to what extent has your physical health or emotional problems interfered with your normal social activities with family, friends, neighbours or group?  Not at all | sf20 | SF-36 |
|  |  |  | During the past 4 weeks, how much of the time has your physical health or emotional problems interfered with your social activities (like visiting friends, relatives, etc.)?  All of the time | sf32 | SF-36 |
|  | Access due to physical wellbeing | *[The chest infection] just made it miserable for a week or two, I couldn’t get out or about…[Male, 75], Al-Janabi, N Flynn, and Coast (2012)* | Do I need any help looking after myself?  I need no help at all | aqol1_4D | AQOL-4D |
|  |  |  | When doing household tasks: (For example: preparing food, gardening, using the video recorder, radio, telephone or washing the car.)  I need no help at all | aqol2_4D | AQOL-4D |
|  |  |  | Thinking about your health and your role in your community (that is to say neighbourhood, sporting, work, church or cultural groups):  My role in the community is unaffected by my health | aqol4 | AQOL-8D |
|  |  |  | Thinking about your health and your relationship with your family:  My role in my family is unaffected by my health | aqol9 | AQOL-8D |
|  |  |  | Thinking about washing yourself, toileting, dressing, eating or looking after your appearance:  These tasks are very easy for me | aqol19 | AQOL-8D |
|  |  |  | How often does pain interfere with your usual activities?  Never | aqol24 | AQOL-8D |
|  |  |  | How much help do you need with tasks around the house (e.g., preparing food, cleaning the house or gardening):  I can do all these tasks very quickly and efficiently without any help | aqol30 | AQOL-8D |
|  |  |  | During the past 4 weeks, have you had any of the following problems with your work or other regular daily activities as a result of your PHYSICAL health? Cut down the amount of time you spent on work or other activities.  All of the time | sf13 | SF-36 |
|  |  |  | During the past 4 weeks, have you had any of the following problems with your work or other regular daily activities as a result of your PHYSICAL health? accomplished less than you would like  All of the time | sf14 | SF-36 |
|  |  |  | During the past 4 weeks, have you had any of the following problems with your work or other regular daily activities as a result of your PHYSICAL health? were limited in the kind of work or other activities  All of the time | sf15 | SF-36 |
|  |  |  | During the past 4 weeks, have you had any of the following problems with your work or other regular daily activities as a result of your PHYSICAL health? had difficulty performing work or other activities (for example, it took extra effort)  All of the time | sf16 | SF-36 |
|  |  |  | During the past 4 weeks, how much did pain interfere with your normal work (including both work outside the home and housework)?  Not at all | sf22 | SF-36 |
|  | Access due to emotional wellbeing | *I started getting depression… it’s like yesterday, I didn’t have a wash, I didn’t have a shave, I didn’t get up, I didn’t even unlock the door, and that was it (Male, not employed, A), Kinghorn, Robinson, and Smith (2015)* | During the past 4 weeks, have you had any of the following problems with your work or other regular daily activities as a result of your EMOTIONAL problems (such as feeling depressed or anxious)?  Cut down the amount of time you spent on work or other activities  All of the time. | sf17 | SF-36 |
|  |  |  | During the past 4 weeks, have you had any of the following problems with your work or other regular daily activities as a result of your EMOTIONAL problems (such as feeling depressed or anxious)?  accomplished less than you would like  All of the time | sf18 | SF-36 |
|  |  |  | During the past 4 weeks, have you had any of the following problems with your work or other regular daily activities as a result of your EMOTIONAL problems (such as feeling depressed or anxious)?  didn't do work or other activities as carefully as usual  All of the time | sf19 | SF-36 |
|  | Access due to social wellbeing | *When I came out of hospital he [husband] done everything I mean, he cooked the food and he’s never cooked in his life [laugh] … And all the washing, ironing he did. (Female, 72 years, PC), Sutton and Coast (2014)* | No relevant items |  |  |
|  | Access due to environmental wellbeing | *“I went in for this flat because it’s wheelchair friendly … I’m hoping that I’d lay here in a box, because it was a very deliberate act of me to look for somewhere where I can be independent for as long as possible.” (Female, 67 years, GP), Sutton and Coast (2014)* | Thinking about how easily I can get around my home and community:  I get around my home and community by myself without any difficulty | aqol3_4d | AQOL-4D |
|  |  |  | Thinking about how easy or difficult it is for you to get around by yourself outside your house (eg shopping, visiting):  Getting around is enjoyable and easy | aqol3 | AQOL-8D |
|  | Access due to activity wellbeing | *“Because of the arthritis and that, I can’t work, so there’s… work mates, you know, no Friday night when you’ve got the wages and you can enjoy it, a couple of beers. There’s none of that.” (Male, Not employed), Kinghorn, Robinson, and Smith (2015)* | No relevant items |  |  |
|  | Access due to finances | *[Regarding the choice to do a NIPT] “If I had to pay for it, I would borrow from my friends or relatives. But I would just do anything possible to avoid a miscarriage.” Kibel and Vanstone (2017)* | No relevant items |  |  |
|  |  | *“I’m reasonably fortunate… in so far as that we’ve got the two pensions… we’re able to go off... We grabbed a cheapie flight at the end of April... flew down to Nice...” (male, aged 70), Grewal et al. (2006)* |  |  |  |
|  | Access due to technology | *“I think the insulin pump is fantastic. Because it gives me freedom.” (#24; Woman, 64 years old, Type 1 DM), Engström et al. (2016)* | Thinking about your mobility, including using any aids or equipment such as wheelchairs, frames, sticks:  I am very mobile | aqol15 | AQOL-8D |
| Perceived control | Management | *“This morning, I got up—5 o’clock—I took my first pain killers, went back to bed again so that I was ready to get up to have my shower at half past six, or else, by the time you start taking them they haven’t taken effect and you’re trying to move around. So, yeah, you’ve got to think ahead…” (Female, not employed, A), Kinghorn, Robinson, and Smith (2015)* | How much do you feel you can cope with life’s problems?  Completely | aqol21 | AQOL-8D |
|  |  | *[Regarding the management of diabetes] “It’s not easy, it’s an endless struggle to try to maintain good blood glucose levels. (...) It’s like walking a line.” (#24; Woman, 64 years old, Type 1 DM), Engström et al. (2016)* |  |  |  |
|  | Evaluation | *It is a constant sadness, that I’ve lost my sight... (...) But it’s nothing I get hung up on in my everyday life. (...) I consider myself as having a good quality of life. (#1; Man, 49 years old, Type 1 DM)* | How often do you feel in control of your life?  Always | aqol29 |  |
|  |  |  | How much of a burden do you feel you are to other people?  Not at all | aqol26 |  |
|  |  |  | And still thinking about the last seven days: how often did you feel worried:  Never | aqol18 |  |
| Option Wellbeing | Item directly linked to theme |  | I am satisfied with my life | SWLS_c | SWLS |
|  |  |  | Overall, how satisfied are you with your life nowadays | ONSi | IHS |
|  |  |  | Thinking about your own life and personal circumstances, how satisfied are you with your life as a whole? | PWI_a | PWI |
|  | Physical wellbeing | *“I just wouldn’t want to be in pain all the time.“ (Female, 72 years, PC), Sutton and Coast (2014)* | How satisfied are you with your health? | PWI_c | PWI |
|  | Emotional wellbeing | *It’s sad not daring to go [on a trip]. (...) Since it [hypo- glycaemia] is a threat, it feels like a lower quality of life. (...) You get a little scared of exposing yourself to situations other than what you are used to. (#17; Woman, 60 years old, Type 2 DM), Engström et al. (2016)* | How often do you feel pleasure?  Always | aqol25 | AQOL-8D |
|  |  |  | How often do you feel happy?  All the time | aqol20 | AQOL-8D |
|  |  |  | How often do you feel sad?  Never | aqol5 | AQOL-8D |
|  |  |  | How content are you with your life?  Extremely | aqol27 | AQOL-8D |
|  |  |  | How often do you feel depressed?  Never | aqol33 | AQOL-8D |
|  |  |  | How often did you feel in despair over the last seven days?  Never | aqol35 | AQOL-8D |
|  |  |  | How much of the time during the past 4 weeks …  Have you felt down  All the time | sf28 | SF-36 |
|  |  |  | How much of the time during the past 4 weeks …  Did you feel worn out  All the time | sf29 | SF-36 |
|  |  |  | How much of the time during the past 4 weeks …  Have you been a happy person  All the time | sf30 | SF-36 |
|  |  |  | How much of the time during the past 4 weeks …  Have you been a very nervous person  All the time | sf24 | SF-36 |
|  |  |  | How much of the time during the past 4 weeks …  Have you felt so down in the dumps that nothing could cheer you up  All the time | sf25 | SF-36 |
|  |  |  | In general, would you say your health is  Excellent | sf1 | SF-36 |
|  |  |  | Overall, how anxious did you feel yesterday? | ONSl | IHS |
|  |  |  | Overall, how happy did you feel yesterday? | ONSk | IHS |
|  |  |  | Enjoyment and pleasure  I can have a lot of enjoyment and pleasure | ic05 | ICECAP - A |
|  |  |  | Thinking about how I generally feel:  I do not feel anxious, worried or depressed | aqol11_4D | AQOL-4D |
|  | Social wellbeing | *“One should take good care of the kids and the entire family, so that everyone is healthy and they can work properly and prosper." Greco et al. (2015)* | Thinking about my relationship with other people:  o I have plenty of friends, and am never lonely | aqol5_4D | AQOL-4D |
|  |  |  | Your close and intimate relationships (including any sexual relationships) make you feel:  o Very happy | aqol34 | AQOL-8D |
|  |  |  | How satisfied are you with your personal relationships? | PWI_e | PWI |
|  |  |  | Your close relationships (family and friends) are:  Very satisfying | aqol10 | AQOL-8D |
|  |  |  | How much do you enjoy your close relationships (family and friends)?  Immensely | aqol23 | AQOL-8D |
|  | Environmental wellbeing | *“A house should have a toilet, a bathing shelter, there should be a rubbish pit, and the house should be well taken care of. Even if you have all these things but they are not put to good use, diseases will be there.” Greco et al. (2015)* | How satisfied are you with feeling part of your community? | PWI_g | PWI |
|  | Activity wellbeing | *“Work is important. Just to go out and do things that aren’t mind numbing if you know what I mean.” (F employed), Kinghorn, Robinson, and Smith (2015)* | No relevant items |  |  |
| Self-Realization | Having a role | *“I do like playing…competitive sport…it’s got a bit of an edge …. I suppose through that there’s a bit of an achievement thing and it’s quite nice to be in a team or to be a captain for one of the teams” (Male, 29), Al-Janabi, N Flynn, and Coast (2012)* | How satisfied are you with what you are achieving in life? | PWI_d | PWI |
|  |  |  | Overall, to what extent do you feel that the things you do in your life are worthwhile? | ONSj | IHS |
|  |  |  | Achievement and progress  I can achieve and progress in all aspects of my life | ic04 | ICECAP - A |
|  | Having dignity | *“A person who changes clothes is seen as living a good life. She changes dirty clothes after a bath, and puts on clean ones, and looks good. When she is amongst people, she is not shy. As for me, I may have to wash the few I have to put on when I go in public.” Greco et al. (2015)* | No relevant items |  |  |
|  | Being independent | *“I would like to drive a bit more. Because I’m losing my independence. I have to rely on my husband to take me shopping now.” (Female, Retired), Kinghorn, Robinson, and Smith (2015)* | Being independent  I am able to be completely independent | ic03 | ICECAP - A |
|  |  | *“A person should be independent because when sick she doesn’t wait for someone to tell her what to do, men at times neglect that you are struggling.” Greco et al. (2015)* |  |  |  |
|  | Self-determination | *[Regarding the choice of conducting NIPT] “I just really think that women should be given ownership of the information and they can decide what they want to do.” Kibel and Vanstone (2017)* | In most ways my life is close to my ideal  Strongly Agree | SWLS_a | SWLS |
|  |  |  | So far I have gotten the important things I want in life  Strongly Agree | SWLS_d | SWLS |

*Appendix Table A2. Standardized factor loadings per factor of model 5.*

|  | Reflective Wellbeing | Affective Wellbeing | Perceived Access to Options | Perceived Control | Social Aspects | Need for Help or Support | Anxiety | Happiness | SF-36 Negative Emotions | SF-Phys Testlet | SF-Emo Testlet |
| --- | --- | --- | --- | --- | --- | --- | --- | --- | --- | --- | --- |
| PWI_a | 0.868 |  |  |  |  |  |  |  |  |  |  |
| PWI_c | 0.454 |  | 0.406 |  |  |  |  |  |  |  |  |
| PWI_d | 0.826 |  |  |  |  |  |  |  |  |  |  |
| PWI_e | 0.638 |  |  |  | 0.327 |  |  |  |  |  |  |
| PWI_g 5 | 0.623 |  |  |  |  |  |  |  |  |  |  |
| ONSi | 0.929 |  |  |  |  |  |  |  |  |  |  |
| ONSj | 0.817 |  |  |  |  |  |  |  |  |  |  |
| SWLS_a | 0.861 |  |  |  |  |  |  |  |  |  |  |
| SWLS_c | 0.910 |  |  |  |  |  |  |  |  |  |  |
| SWLS_d | 0.793 |  |  |  |  |  |  |  |  |  |  |
| aqol27 | 0.406 | 0.524 |  |  |  |  |  | 0.152 |  |  |  |
| sf1 | 0.293 |  | 0.544 |  |  |  |  |  |  |  |  |
| ONSl |  | 0.446 |  |  |  |  | 0.399 |  |  |  |  |
| sf17 |  | 0.436 | 0.310 |  |  |  |  |  |  |  | 0.672 |
| sf18 5 |  | 0.495 | 0.263 |  |  |  |  |  |  |  | 0.640 |
| sf19 |  | 0.430 | 0.303 |  |  |  |  |  |  |  | 0.647 |
| sf20 |  | 0.437 | 0.424 |  |  |  |  |  |  |  |  |
| sf24 |  | 0.634 |  |  |  |  | 0.410 |  | 0.335 |  |  |
| sf25 |  | 0.754 |  |  |  |  |  |  | 0.439 |  |  |
| sf28 5 |  | 0.832 |  |  |  |  |  |  | 0.328 |  |  |
| sf29 |  | 0.493 | 0.249 |  |  |  |  |  | 0.221 |  |  |
| sf30 |  | 0.717 |  |  |  |  |  | 0.275 |  |  |  |
| sf32 |  | 0.412 | 0.447 |  |  |  |  |  |  |  |  |
| aqol5 |  | 0.855 |  |  |  |  |  |  |  |  |  |
| aqol9 |  | 0.246 | 0.585 |  |  |  |  |  |  |  |  |
| aqol10 |  | 0.634 |  |  | 0.373 |  |  |  |  |  |  |
| aqol18 |  | 0.787 |  |  |  |  |  |  |  |  |  |
| aqol20 |  | 0.806 |  |  |  |  |  | 0.372 |  |  |  |
| aqol23 |  | 0.561 |  |  | 0.582 |  |  |  |  |  |  |
| aqol25 |  | 0.687 |  |  |  |  |  | 0.152 |  |  |  |
| aqol33 |  | 0.899 |  |  |  |  |  |  |  |  |  |
| aqol34 |  | 0.582 |  |  | 0.373 |  |  |  |  |  |  |
| aqol35 |  | 0.820 |  |  |  |  |  |  |  |  |  |
| aqol5_4D |  | 0.683 |  |  | 0.210 |  |  |  |  |  |  |
| aqol11_4D |  | 0.846 |  |  |  |  | 0.170 |  |  |  |  |
| aqol1_4D |  |  | 0.592 |  |  | 0.715 |  |  |  |  |  |
| aqol2_4D |  |  | 0.732 |  |  |  |  |  |  |  |  |
| aqol3_4D |  |  | 0.646 |  |  | 0.289 |  |  |  |  |  |
| aqol3 |  |  | 0.835 |  |  |  |  |  |  |  |  |
| aqol4 5 |  |  | 0.770 |  |  |  |  |  |  |  |  |
| aqol15 |  |  | 0.793 |  |  |  |  |  |  |  |  |
| aqol19 |  |  | 0.769 |  |  | 0.174 |  |  |  |  |  |
| aqol24 |  |  | 0.748 |  |  |  |  |  |  |  |  |
| aqol26 |  |  | 0.272 | 0.550 |  |  |  |  |  |  |  |
| aqol30 |  |  | 0.829 |  |  |  |  |  |  |  |  |
| sf13 |  |  | 0.694 |  |  |  |  |  |  | 0.544 |  |
| sf14 |  |  | 0.712 |  |  |  |  |  |  | 0.547 |  |
| sf15 |  |  | 0.763 |  |  |  |  |  |  | 0.566 |  |
| sf16 |  |  | 0.768 |  |  |  |  |  |  | 0.566 |  |
| sf22 |  |  | 0.754 |  |  |  |  |  |  |  |  |
| ic03 |  |  | 0.377 | 0.312 |  |  |  |  |  |  |  |
| aqol21 |  |  |  | 0.813 |  |  |  |  |  |  |  |
| aqol29 |  |  |  | 0.837 |  |  |  |  |  |  |  |
| ic04 |  |  |  | 0.746 |  |  |  |  |  |  |  |
| ic05 |  |  |  | 0.548 |  |  |  |  |  |  |  |

*Appendix Table 3. Item error correlations of model 5.*

| Items | Error Correlation value |
| --- | --- |
| PWI_c – sf1 | 0.496 |
| aqol24 – sf22 | 0.489 |
| sf20 – sf32 | 0.444 |
| aqol2_4D – aqol30 | 0.302 |

*Appendix Table 4. Standardized inter-factor correlations of model 5.*

| Factors | Reflective Wellbeing | Affective Wellbeing | Perceived Control | Perceived Access to Options | Physical limitations testlet | Emotional limitations testlet |
| --- | --- | --- | --- | --- | --- | --- |
| Reflective Wellbeing | 1 |  | | |  |  |
| Affective Wellbeing | 0.755 | 1 |  | |  |  |
| Perceived Control | 0.772 | 0.913 | 1 |  |  |  |
| Perceived Access to Options | 0.406 | 0.530 | 0.615 | 1 |  |  |
| Physical limitations teslet | 0 | 0 | 0 | 0 | 1 |  |
| Emotional limitations teslet | 0 | 0 | 0 | 0 | 0.462 | 1 |

*Appendix Table 5. Proportion of responses per option of the WeRFree instrument.*

| Items and response option level | Percentage of sample choosing specific response option |
| --- | --- |
| PWI_a |  |
| 1 | 1.9% |
| 2 | 1.9% |
| 3 | 4.6% |
| 4 | 6.0% |
| 5 | 6.3% |
| 6 | 10.7% |
| 7 | 10.3% |
| 8 | 18.9% |
| 9 | 23.3% |
| 10 | 0.9% |
| 11 | 5.3% |
| PWI_e |  |
| 1 | 2.9% |
| 2 | 3.0% |
| 3 | 3.9% |
| 4 | 4.4% |
| 5 | 4.7% |
| 6 | 10.6% |
| 7 | 07.5% |
| 8 | 11.9% |
| 9 | 16.6% |
| 10 | 18.1% |
| 11 | 16.4% |
| PWI_d |  |
| 1 | 3.0% |
| 2 | 3.4% |
| 3 | 4.8% |
| 4 | 5.7% |
| 5 | 7.3% |
| 6 | 13.3% |
| 7 | 11.1% |
| 8 | 16.2% |
| 9 | 17.7% |
| 10 | 11.7% |
| 11 | 5.9% |
| PWI_g |  |
| 1 | 2.5% |
| 2 | 2.6% |
| 3 | 3.7% |
| 4 | 3.9% |
| 5 | 5.0% |
| 6 | 19.7% |
| 7 | 10.0% |
| 8 | 14.8% |
| 9 | 17.7% |
| 10 | 11.9% |
| 11 | 7.9% |
| ONSj |  |
| 1 | 2.2% |
| 2 | 3.1% |
| 3 | 3.7% |
| 4 | 3.8% |
| 5 | 4.6% |
| 6 | 12.2% |
| 7 | 12.2% |
| 8 | 16.2% |
| 9 | 19.3% |
| 10 | 14.6% |
| 11 | 8.1% |
| SWLS_a |  |
| 1 | 6.7% |
| 2 | 13.6% |
| 3 | 13.0% |
| 4 | 12.3% |
| 5 | 25.3% |
| 6 | 25.0% |
| 7 | 4.1% |
| SWLS_d |  |
| 1 | 4.6% |
| 2 | 8.5% |
| 3 | 11.2% |
| 4 | 12.0% |
| 5 | 22.3% |
| 6 | 30.7% |
| 7 | 10.6% |
| aqol5_4D |  |
| 1 | 5.9% |
| 2 | 16.0% |
| 3 | 36.6% |
| 4 | 41.5% |
| aqol5 |  |
| 1 | 3.8% |
| 2 | 9.0% |
| 3 | 40.0% |
| 4 | 40.8% |
| 5 | 6.4% |
| aqol25 |  |
| 1 | 1.1% |
| 2 | 7.8% |
| 3 | 32.5% |
| 4 | 47.7% |
| 5 | 10.8% |
| aqol35 |  |
| 1 | 1.2% |
| 2 | 7.0% |
| 3 | 14.0% |
| 4 | 22.0% |
| 5 | 55.8% |
| ic05 |  |
| 1 | 1.6% |
| 2 | 21.7% |
| 3 | 42.6% |
| 4 | 34.2% |
| sf24 |  |
| 1 | 2.1% |
| 2 | 7.5% |
| 3 | 19.9% |
| 4 | 31.3% |
| 5 | 39.3% |
| sf30 |  |
| 1 | 5.0% |
| 2 | 15.7% |
| 3 | 25.2% |
| 4 | 43.6% |
| 5 | 10.6% |
| aqol3 |  |
| 1 | 1.3% |
| 2 | 4.3% |
| 3 | 8.2% |
| 4 | 15.4% |
| 5 | 34.9% |
| 6 | 35.9% |
| aqol4 |  |
| 1 | 5.1% |
| 2 | 9.2% |
| 3 | 18.4% |
| 4 | 67.3% |
| aqol9 |  |
| 1 | 1.5% |
| 2 | 8.1% |
| 3 | 26.0% |
| 4 | 64.4% |
| aqol15 |  |
| 1 | 0.0% |
| 2 | 1.2% |
| 3 | 11.7% |
| 4 | 16.7% |
| 5 | 17.4% |
| 6 | 53.0% |
| aqol19 |  |
| 1 | 0.1% |
| 2 | 1.9% |
| 3 | 11.4% |
| 4 | 20.7% |
| 5 | 65.8% |
| aqol24 |  |
| 1 | 3.5% |
| 2 | 10.5% |
| 3 | 23.9% |
| 4 | 33.2% |
| 5 | 28.9% |
| aqol30 |  |
| 1 | 0.8% |
| 2 | 6.0% |
| 3 | 15.0% |
| 4 | 30.9% |
| 5 | 47.3% |

*Appendix Table 6. Standardized factor loadings, intercepts and variances and inter-factor correlations of the WeRFree instrument.*

| Construct  Item | Standardized loadings | Standardized Intercepts | Standardized Variances |
| --- | --- | --- | --- |
| Reflective Wellbeing |  |  |  |
| PWI_a | 0.851 | 3.155 | 0.276 |
| PWI_e | 0.693 | 2.928 | 0.520 |
| PWI_g | 0.661 | 2.964 | 0.563 |
| ONSj | 0.824 | 3.011 | 0.322 |
| SWLS_a | 0.839 | 2.602 | 0.295 |
| SWLS_d | 0.785 | 2.915 | 0.385 |
| Affective Wellbeing |  |  |  |
| sf24 | 0.627 | 3.926 | 0.607 |
| sf30 | 0.725 | 3.335 | 0.459 |
| aqol5 | 0.830 | 3.846 | 0.312 |
| aqol35 | 0.813 | 4.193 | 0.340 |
| Perceived Access to Options |  |  |  |
| aqol3 | 0.848 | 4.265 | 0.281 |
| aqol4 | 0.764 | 4.177 | 0.431 |
| aqol19 | 0.788 | 6.015 | 0.380 |
| aqol24 | 0.743 | 3.503 | 0.448 |
| aqol30 | 0.812 | 4.552 | 0.341 |
| Standardized factor correlations | | | |
|  | Reflective Wellbeing | Affective Wellbeing | Perceived Access to Options |
| Reflective wellbeing | 1 | - | - |
| Affective wellbeing | 0.762 | 1 | - |
| Perceived access to options | 0.406 | 0.560 | 1 |

*Appendix table 7. Item – total correlations per scale of the WeRFree instrument*

| Item | Item-total correlation |
| --- | --- |
| PWI_a | 0.79 |
| PWI_e | 0.67 |
| PWI_g | 0.64 |
| ONSj | 0.77 |
| SWLS_a | 0.77 |
| SWLS_d | 0.72 |
| sf24 | 0.58 |
| sf30 | 0.62 |
| aqol5 | 0.74 |
| aqol35 | 0.73 |
| aqo3 | 0.78 |
| aqol4 | 0.70 |
| aqol19 | 0.73 |
| aqol24 | 0.70 |
| aqol30 | 0.76 |

### Appendix Section 3. Items included in the WeRFree instrument.

| **Reflective Wellbeing** | | | | | | | | | | | | | | | | | | | | | | |  |
| --- | --- | --- | --- | --- | --- | --- | --- | --- | --- | --- | --- | --- | --- | --- | --- | --- | --- | --- | --- | --- | --- | --- | --- |
|  | Completely dissatisfied |  | | |  | | |  | | |  | | | Neutral | | |  | |  | |  |  | Completely satisfied |
|  | 0 | 1 | | | 2 | | | 3 | | | 4 | | | 5 | | | 6 | | 7 | | 8 | 9 | 10 |
| Thinking about your own life and personal circumstances, how satisfied are you with your life as a whole? |  |  | | |  | | |  | | |  | | |  | | |  | |  | |  |  |  |
| How satisfied are you with your personal relationships? |  |  | | |  | | |  | | |  | | |  | | |  | |  | |  |  |  |
| How satisfied are you with feeling part of your community? |  |  | | |  | | |  | | |  | | |  | | |  | |  | |  |  |  |
|  | Not at all Worthwhile |  | | |  | | |  | | |  | | | Neutral | | |  | |  | |  |  | Completely worthwhile |
| Overall, to what extent do you feel that the things you do in your life are worthwhile? |  |  | | |  | | |  | | |  | | |  | | |  | |  | |  |  |  |
|  | Strongly disagree | | Disagree | | | Slightly disagree | | | neither agree nor disagree | | | slightly agree | | | agree | | | strongly agree | |  |  |  |  |
| In most ways my life is close to my ideal: |  | |  | | |  | | |  | | |  | | |  | | |  | |  |  |  |  |
| So far I have gotten the important things I want in life: |  | |  | | |  | | |  | | |  | | |  | | |  | |  |  |  |  |
| **Affective Wellbeing** | | | | | | | | | | | | | | | | | | | |  |  |  |  |
|  | All of the time | | | Most of the time | | | Some of the time | | | A little of the time | | | None of the time | | |  |  |  |  |  |  |  |  |
| How much of the time during the past 4 weeks have you been a very nervous person? |  | | |  | | |  | | |  | | |  | | |  |  |  |  |  |  |  |  |
|  | All of the time | | | Most of the time | | | Some of the time | | | A little of the time | | | None of the time | | |  |  |  |  |  |  |  |  |
| How much of the time during the past 4 weeks have you been a happy person? |  | | |  | | |  | | |  | | |  | | |  |  |  |  |  |  |  |  |
| How often do you feel sad? | | | | | | | | | | | | | | | | | | | | | | | |
| O Never  O Rarely  O Some of the time  O Usually  O Nearly all the time | | | | | | | | | | | | | | | | | | | | | | | |
| How often did you feel in despair over the last seven days? | | | | | | | | | | | | | | | | | | | | | | | |
| O Never  O Occasionally  O Sometimes  O Often  O All the time  **Perceived Access to Options** | | | | | | | | | | | | | | | | | | | | | | | |
| Thinking about how easy or difficult it is for you to get around by yourself outside your house (e.g. shopping, visiting): | | | | | | | | | | | | | | | | | | | | | | | |
| O Getting around is enjoyable and easy  O I have no difficulty getting around outside my house  O A little difficulty  O Moderate difficulty  O A lot of difficulty  O I cannot get around unless somebody is there to help me | | | | | | | | | | | | | | | | | | | | | | | |
| Thinking about your health and your role in your community (that is to say neighborhood, sporting, work, church or cultural groups): | | | | | | | | | | | | | | | | | | | | | | | |
| O My role in the community is unaffected by my health  O There are some parts of my community role I cannot carry out  O There are many parts of my community role I cannot carry out  O I cannot carry out any part of my community role | | | | | | | | | | | | | | | | | | | | | | | |
| Thinking about washing yourself, toileting, dressing, eating or looking after your appearance: | | | | | | | | | | | | | | | | | | | | | | | |
| O These tasks are very easy for me  O I have no real difficulty in carrying out these tasks  O I find some of these tasks difficult, but I manage to do them on my own  O Many of these tasks are difficult, and I need help to do them  O I cannot do these tasks by myself at all | | | | | | | | | | | | | | | | | | | | | | | |
| How often does pain interfere with your usual activities? | | | | | | | | | | | | | | | | | | | | | | | |
| O Never  O Almost never  O Sometimes  O Often  O Always | | | | | | | | | | | | | | | | | | | | | | | |
| How much help do you need with tasks around the house (eg preparing food, cleaning the house or gardening): | | | | | | | | | | | | | | | | | | | | | | | |
| O I can do all these tasks very quickly and efficiently without any help  O I can do these tasks relatively easily without help  O I can do all these tasks only very slowly without help  O I cannot do most of these tasks unless I have help  O I can do none of these tasks by myself | | | | | | | | | | | | | | | | | | | | | | | |

Al-Janabi, Hareth, Terry N Flynn, and Joanna Coast. 2012. "Development of a self-report measure of capability wellbeing for adults: the ICECAP-A." *Quality of Life Research* 21 (1):167-76. doi: 10.1007/s11136-011-9927-2.

Bandura, Albert. 2001. "Social cognitive theory: An agentic perspective." *Annual review of psychology* 52 (1):1-26.

Brazier, John, Jennifer Roberts, and Mark Deverill. 2002. "The estimation of a preference-based measure of health from the SF-36." *Journal of health economics* 21 (2):271-92.

Cummins, Robert A, Richard Eckersley, Julie Pallant, Jackie Van Vugt, and RoseAnne Misajon. 2003. "Developing a national index of subjective wellbeing: The Australian Unity Wellbeing Index." *Social indicators research* 64 (2):159-90.

Diener, ED, Robert A Emmons, Randy J Larsen, and Sharon Griffin. 1985. "The satisfaction with life scale." *Journal of personality assessment* 49 (1):71-5.

Dolan, Paul, and Robert Metcalfe. 2012. "Measuring subjective wellbeing: Recommendations on measures for use by national governments." *Journal of social policy* 41 (2):409-27.

Engström, Svedbo, Maria Leksell, Janeth Johansson, Unn-Britt, and Gudbjörnsdottir Soffia. 2016. "What is important for you? A qualitative interview study of living with diabetes and experiences of diabetes care to establish a basis for a tailored Patient-Reported Outcome Measure for the Swedish National Diabetes Register." *BMJ Open* 6 (3). doi: 10.1136/bmjopen-2015-010249.

Frei, Anja, Anna Svarin, Claudia Steurer-Stey, and Milo A Puhan. 2009. "Self-efficacy instruments for patients with chronic diseases suffer from methodological limitations-a systematic review." *Health and quality of life outcomes* 7 (1):1-10.

Greco, Giulia, Jolene Skordis-Worrall, Bryan Mkandawire, and Anne Mills. 2015. "What is a good life? Selecting capabilities to assess women's quality of life in rural Malawi." *Social Science & Medicine* 130:69-78. doi: <https://doi.org/10.1016/j.socscimed.2015.01.042>.

Grewal, Ini, Jane Lewis, Terry Flynn, Jackie Brown, John Bond, and Joanna Coast. 2006. "Developing attributes for a generic quality of life measure for older people: Preferences or capabilities?" *Social Science & Medicine* 62 (8):1891-901. doi: <https://doi.org/10.1016/j.socscimed.2005.08.023>.

Hawthorne, Graeme, Jeff Richardson, and Richard Osborne. 1999. "The Assessment of Quality of Life (AQoL) instrument: a psychometric measure of health-related quality of life." *Quality of Life Research* 8 (3):209-24.

Kibel, Mia, and Meredith Vanstone. 2017. "Reconciling ethical and economic conceptions of value in health policy using the capabilities approach: a qualitative investigation of non-invasive prenatal testing." *Social Science & Medicine* 195:97-104.

Kinghorn, Philip, Angela Robinson, and Richard D. Smith. 2015. "Developing a Capability-Based Questionnaire for Assessing Well-Being in Patients with Chronic Pain." *Social indicators research* 120 (3):897-916. doi: 10.1007/s11205-014-0625-7.

Richardson, J, MA Khan, A Iezzi, and A Maxwell. 2012. "Cross-national comparison of twelve quality of life instruments. Research papers 78, 80–83, 85." *MIC report* 2.

Richardson, Jeff, Munir Khan, Angelo Iezzi, Kompal Sinha, Cathy Mihalopoulos, Helen Herrman, Graeme Hawthorne, and Issy Schweitzer. 2009. "The AQoL-8D (PsyQoL) MAU Instrument: Overview September 2009." *Centre for Health Economics, Monash University*.

Seiber, William J, Erik J Groessl, Kristin M David, Theodore G Ganiats, and Robert M Kaplan. 2008. "Quality of well being self-administered (QWB-SA) scale."

Sutton, Eileen J, and Joanna Coast. 2014. "Development of a supportive care measure for economic evaluation of end-of-life care using qualitative methods." *Palliative Medicine* 28 (2):151-7. doi: 10.1177/0269216313489368.
